# Maternal blood and amnionic oxytocin receptor gene expression and serum oxytocin levels in preterm birth: a case control study

**DOI:** 10.1101/2020.07.18.20155879

**Authors:** Kumari Anukriti, Kiran Guleria, Vipin Tyagi, Amita Suneja, B D Banerjee

## Abstract

**Background:** The oxytocin (OXT)-oxytocin receptor (*OXTR)* system provides promising candidate gene for studies of genetic contributions to prematurity.

**Objective:** Quantification and comparison of oxytocin receptor (*OXTR*) gene expression and serum OXT levels in the blood and amnion of women delivering preterm and evaluation of the correlation between *OXTR* gene expression in blood and amnion with serum OXT levels in them.

**Methods:** 70 pregnant women in spontaneous labor delivering vaginally preterm i.e < 37 weeks and equal number of matched controls delivering spontaneously at term (37-42 weeks) were recruited. Maternal serum OXT levels taken in active stage of labor i.e 4 cm cervical dilatation were quantified by ELISA. Gene expression studies in the maternal blood and amnion were done by using real time quantitative polymerase chain reaction (RT-qPCR).

**Results:** The mean serum OXT level in PTL was 48.56 ± 6.97 pg/ml; significantly higher than in controls (43.00 ± 3.96 pg/ml), p<0.001. OXTR gene expression both in maternal blood (2.5 times) and amnion (3.5 times) were significantly higher in PTL. A significant positive correlation was observed between serum OXT levels and OXTR gene expression in amnion (r = −0.190, p = 0.025).

**Conclusions:** The serum OXT levels and OXTR gene expression in amnion surge significantly in active phase of PTL. Thus, amnion probably links OXT-PTGs autocrine paracrine circuit to facilitate PTL. Future studies are needed to devise better OXTR receptor antagonists preferably acting on amnionic OXTRs to prevent PTL.

## Introduction

An estimated 15 million babies are born too early every year i.e. 1 in 10 babies. Approximately 1 million children die each year due to complications of preterm birth (PTB) [1]. Globally, prematurity is the leading cause of death in children under the age of 5 years. Many survivors face a lifetime of disability, including learning disabilities, visual and hearing problems. PTB rates are increasing despite advancing knowledge of risk factors for preterm labour (PTL) and the introduction of public health and medical interventions. Key treatments for PTL have focused on the prevention or inhibition of myometrial contractions, mainly to provide time to administer steroids to aid fetal lung maturation and transfer to a special neonatal care unit [2,3]. Tocolytics currently in use have limited effectiveness and on average only delay labour by up to 48 hours. Understanding more about the mechanisms of labour is essential to identify targets for novel and more effective therapies to stop or prevent PTL. The neurohypophysial hormone oxytocin (OXT) is named after the “quick birth” which it causes due to its uterotonic activity. Oxytocin receptor (OXTR) gene expression has been shown to be present in humans in amnion, chorion, and decidua. OXT binding to OXTR significantly increases in human fetal membranes specially amnion with the onset of labor. Terzidou et al [4] revealed that human OXTR expression increases spontaneously in postlabor amnion epithelial cells and that treatment with interleukin (IL)1B stimulates OXTR expression in prelabor amnion when cultured. The increased ability of human amnion to produce prostaglandin E(PGE) in response to OXT treatment suggests a complementary role of the OXT/OXTR system in the activation of PG pathway in the human amnion resulting in the onset of labor.

OXT stimulates myometrial contractions through multiple signalling pathways. Binding of OXT to its receptor has been known to lead to G-protein coupling and subsequently, increase in intracellular calcium levels to mediate the generation of force [5]. There is a role for the OXT-OXTR system in the onset of human labour, additional to the stimulation of myometrial contractions, that involves increase in the expression of cyclooxygenase-2 (COX-2) and other inflammatory mediators known to be associated with the onset of labour [6,7]. OXT-OXTR interaction leads to NF-κB activation and subsequent upregulation of PGs, inflammatory chemokines and cytokines that are known to play roles in fetal membrane remodelling, cervical ripening and myometrial activation. Amnion is an important site of PG production within the human uterus and its activation is critical for fetal membrane remodelling, cervical ripening and the stimulation of myometrial contraction [6]. Therefore, OXTR is commonly used as a target for the development of tocolytics. *Atosiban*, an OXTR antagonist acting on both myometrial & decidual OXTRs has been used to arrest premature uterine contraction but has failed to reduce the incidence of PTB or improve the neonatal outcome compared with placebo. The role of OXT in PG pathway activation leading to onset of labour suggests that potential clinical use of OXTR antagonists requires re-evaluation. The study of molecular genetics involving OXT-OXTR system in gestational tissues may prove fruitful in deciphering these complex mechanisms leading to PTB. The present study gains insight into OXT-OXTR system in PTB in order to build the ground for future research for better targeted interventions.

## Materials and Methods

### Subjects

In this case-control study, one hundred forty study subjects (n=140) in spontaneous labour were recruited at University College of Medical Sciences (UCMS) & Guru Teg Bahadur Hospital, Delhi, India from November 2014 to April 2016. An informed consent was taken from all the study subjects and ethical approval was taken from the Institutional Ethics Committee for Human Research (IEC-HR), UCMS & GTB hospital, University of Delhi, India. Seventy women aged 18 to 35 years with BMI 19 to 26 kg/m^2^ in spontaneous PTL with intact membranes who delivered vaginally appropriate for gestational age neonates were recruited as cases. Women having history of one or more spontaneous PTB or late second trimester spontaneous abortion or cervical cerclage were excluded from the study. Women with multiple pregnancy, fetal growth restriction, stillbirth, anemia, diabetes, hypertension, chronic diseases, urinary tract infections, chorioamnioitis, recent intake of anti-inflammatory drugs/steroids, history of smoking or any complications during pregnancy and/or in labor were excluded from the study groups. For each case, an age, weight and parity matched, low risk women (n=70) in spontaneous labour delivering vaginally at term (37-42 weeks) appropriate for gestational age neonate were recruited as controls.

### Sample Collection and Storage

Maternal blood (2ml) was collected in an EDTA vial during the active phase of labour (≥4 cm cervical dilatation) and 250 µl of it was fixed in 750 µl of TRIzol LS (Ambion, USA) reagent and stored at −80°C. Maternal serum (2ml) was separated by centrifugation and stored at −20°C. Serum OXT levels were estimated by ELISA using OXT Enzyme Immune Assay Kit (K048-H1) by Arbor Assays (Ann Arbor, Michigan, US) with declared sensitivity as 17.0 pg/mL with limit of detection as 22.9 pg/ml as per manufacturer’s protocol.

Amnion was carefully separated from chorion just after delivery of placenta. A 5×5 cm^2^ of amnion was collected, cut into small strips and washed in sterile phosphate buffered saline (PBS). One gm of homogenized amnion was fixed in 1ml of TRIzol reagent & stored at −80°C.

Ribonucleic acid (RNA) was isolated from maternal blood & amnionic tissue and used for complimentary deoxynucleic acid (cDNA) synthesis using Maxima first strand cDNA synthesis kit (Fermentas, USA). It was used to study *OXTR* gene expression in maternal blood and amnionic tissue by Real Time quantitative polymerase chain reaction (RT-qPCR) using CFX Connect (Biorad, USA) RT-qPCR machine and corresponding ΔCq (Quantification cycle) values were determined by using gene specific primers. The primers sequences used for the amplification of human OXTR and β-actin were following: forward5’ACT TTA GGT TCG CCT GCG G 3’ and reverse 5’CTC CTC TGA GCC ACT GCA AA3’(Primer for OXTR, amplicon size-126bp) and forward 5’TCA CAA GGA TTC CTA TGT GG3’5’CTC ATT GTA GAA GGT GTG G 3’(Primer for β-actin, amplicon size-136bp).

The qPCR amplification master mix for a sample of a gene was made, 20 µl which contained 1 µl of cDNA, 10 µl SSO Fast Evagreen SuperMix, 1 µl each of forward and reverse specific primer pairs (conc. 10 pmol/ µl), 7 µl of nuclease free water. All reactions were set in duplicate along with no template control. The PCR was carried out for initial denaturation (95 ° C for 5min) followed by 40 cycles consisting of template denaturation (10 sec at 94 ° C), primer annealing and extension (40 sec at 60 ° C) and run under appropriat cycling conditions (Table 1).

**Table 1:** Thermoprofile used for (qPCR)

| Gene | Initial Denaturation | Denaturation | Annealing/Extension |
| --- | --- | --- | --- |
| OXTR | 95°C, 5 min | 95°C, 10 sec (40 cycles) | 60°C, 40 sec (40 cycles) |
| B-Actin | 95°C, 5 min | 95°C, 10 sec (40 cycles) | 60°C, 40 sec (40 cycles) |

In the initial cycles of PCR, there is a little change in fluorescence signal (produced from double stranded DNA). This defines the baseline for the amplification plot. An increase in fluorescence above the baseline indicates the detection of accumulated target. The parameter Cq is defined as the fractional cycle number at which the fluorescence passes the fixed threshold. Cq levels are inversely proportional to the amount of target nucleic acid in the sample i.e. lower the value of Cq, higher the amount of target nucleic acid in the sample. Expression normalization was done by β-actin gene to correct for sample to sample variations in qPCR efficiency and errors in sample quantification. ΔCq was evaluated which is the difference of average Cq of target gene (OXTR gene) and reference gene (β-actin gene).

ΔCq = Average Cq_^OXTR^ - Average Cq_ ^β-actin gene^. Again, the difference of mean Cq values of control and cases was determined, which is ΔΔCq.

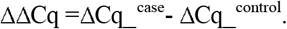

After this, true Fold change (FC) was represented to compare the expression of genes between cases and controls by the following formula: FC= 2 ^−ΔΔCq^

### Statistical analysis

Microsoft Excel (version 2007) and statistical software SSPS for windows (version 17.0) was used for data presentation and statistical analysis. Unpaired Student’s t-test and Chi-square/Fisher’s exact test was applied to compare all socio-demographic characteristics, clinical profile, OXTR gene expression in cases and controls. Pearson coefficient of correlation was used for various correlation studies.

## Results

The comparison of various socio demographic features in the two groups is depicted in Table 2. The mean serum OXT level in active labour in PTB cases was 48.56 ± 6.2 pg/ml which was significantly higher (*p*<0.001) than those with term labour (43.00 ± 3.96 pg/ml) (Table 3). In subgroup analysis of early v/s late preterm, the serum OXT levels continued to be higher in preterm than term but there was no statistically significant difference. The values of OXTR ΔCq in maternal blood of cases and controls are compared in Table 3. The maternal blood OXTR ΔCq value in cases was significantly lower (i.e. expression of OXTR gene in maternal blood was significantly higher) with respect to controls (p <0.001). Also Fold change (FC) was > 2.44 which means its expression was 2.44 times in PTL than that in term labour (Fig 1). OXTR gene expression in amnion was significantly higher in PTL (FC >3.44) as compared to term labour (p<0.001) (Fig 1). However, the value of OXTR ΔCq in maternal blood as well in amnion was not statistically significantly different in early and late preterm subgroups. In correlation studies, a negative correlation was observed between maternal serum OXT levels and OXTR ΔCq in maternal blood (i.e. positive correlation between maternal serum OXT levels and OXTR gene expression in maternal blood) in PTL (Table 4). Similarly, a negative correlation was seen between maternal serum OXT levels and OXTR ΔCq in amnion in PTL (i.e. positive correlation between maternal serum OXT levels and OXTR gene expression in amnion) (Table 4). A negative correlation was observed between maternal serum OXT levels and amnionic OXTR ΔCq (i.e. positive correlation between maternal serum OXT levels and amnionic OXTRs) when all the study subjects (preterm & term) were combined into a single group (Table 5 & Fig. 2). There was a significant proportionate rise in maternal serum OXT levels with increased OXTR gene expression in amnion (p<0.025) in all labouring subjects irrespective of period of gestation.

**Table 2:** Comparison of socio-demographic characteristics of the study groups.

| Characteristics | Group 1<br>Cases (preterm) n=70; Number (%) | Group 2<br>Controls (term) n=70; Number (%) | p value |
| --- | --- | --- | --- |
| Residential Area |  |  |  |
| Slum | 8 (11.4%) | 8 (11.4%) | 0.109 |
| Market | 41 (58.6%) | 33 (47.1%) |  |
| Colony | 21 (46.7%) | 24 (34.3%) |  |
| Industrial Area | 0 (0.0%) | 5 (7.1%) |  |
| Religion |  |  |  |
| Hindu | 53 (75.7%) | 50 (71.4%) | 0.702 |
| Muslim | 17 (24.3%) | 20 (28.6%) |  |
| Socioeconomic status |  |  |  |
| Class I (Upper) | 0 (0.0%) | 0 (0.0%) | 0.033 |
| Class II (Upper-middle) | 2 (2.9%) | 1 (1.4%) |  |
| Class III (Lower-middle) | 37 (52.9%) | 25 (35.7%) |  |
| Class IV (Upper-lower) | 13 (18.6%) | 10 (14.3%) |  |
| Class V (Lower) | 18 (25.7%) | 34 (48.6%) |  |
| Education |  |  |  |
| Illiterate | 11 (15.7%) | 7 (10.0%) | 0.258 |
| Primary | 15 (21.4%) | 9 (12.9%) |  |
| High School | 18 (25.7%) | 15 (21.4%) |  |
| Secondary | 23 (32.9%) | 35 (50.0%) |  |
| Graduate | 3 (4.3%) | 4 (5.7%) |  |
| Occupation |  |  |  |
| Housewife | 67 (95.7%) | 66 (94.3%) | 1.000 |
| Working (ANM / teacher / tailor / beautician) | 3 (4.3%) | 4 (5.7%) |  |
| Dietary habits |  |  |  |
| Vegetarian | 41 (58.6%) | 38 (54.3%) | 0.733 |
| Non-vegetarian | 29 (41.4%) | 32 (45.7%) |  |
| Water source |  |  |  |
| Government supply | 59 (84.3%) | 67 (95.7%) | 0.043 |
| Hand Pump | 4 (5.7%) | 1 (1.4%) |  |
| Well | 5 (3.6%) | 0 (0.0%) |  |
| Pvt. Source | 2 (2.9%) | 2 (2.9%) |  |
| No addiction | 68 (97.1%) | 65 (92.9%) | 0.441 |

p value < 0.05 is significant

**Table 3:** Serum Oxytocin levels in the two study groups and two subgroups:

| Parameter | Group 1<br>Cases<br>(preterm);<br>n=70<br>(mean<br>±SD) | Group 2<br>Controls (term);<br>n=70<br>(mean ±SD) | p-value | Early preterm<br>(< 34 weeks);<br>n=37<br>(mean ± SD) | Late preterm (34-36<sup>+6</sup> weeks); n=33<br>(mean ± SD) | p-value |
| --- | --- | --- | --- | --- | --- | --- |
| Serum<br>Oxytocin<br>levels (pg/ml) | 48.56±6.9<br>7 | 43.00±3.96 | <0.001 | 48.7995 ± 6.68 | 48.2921 ± 7.38 | 0.764 |
| OXTR ΔCq<br>(maternal<br>blood) | 6.400±1.4<br>2 | 7.69±1.47 | <0.001 | 6.26 ± 1.38 | 6.55 ± 1.48 | 0.404 |
| OXTR Δ Cq<br>(Amnion) | 10.88<br>±1.38 | 12.66 ± 2.17 | <0.001 | 10.702 ± 1.46 | 11.088 ± 1.28 | 0.249 |

**Fig 1:**
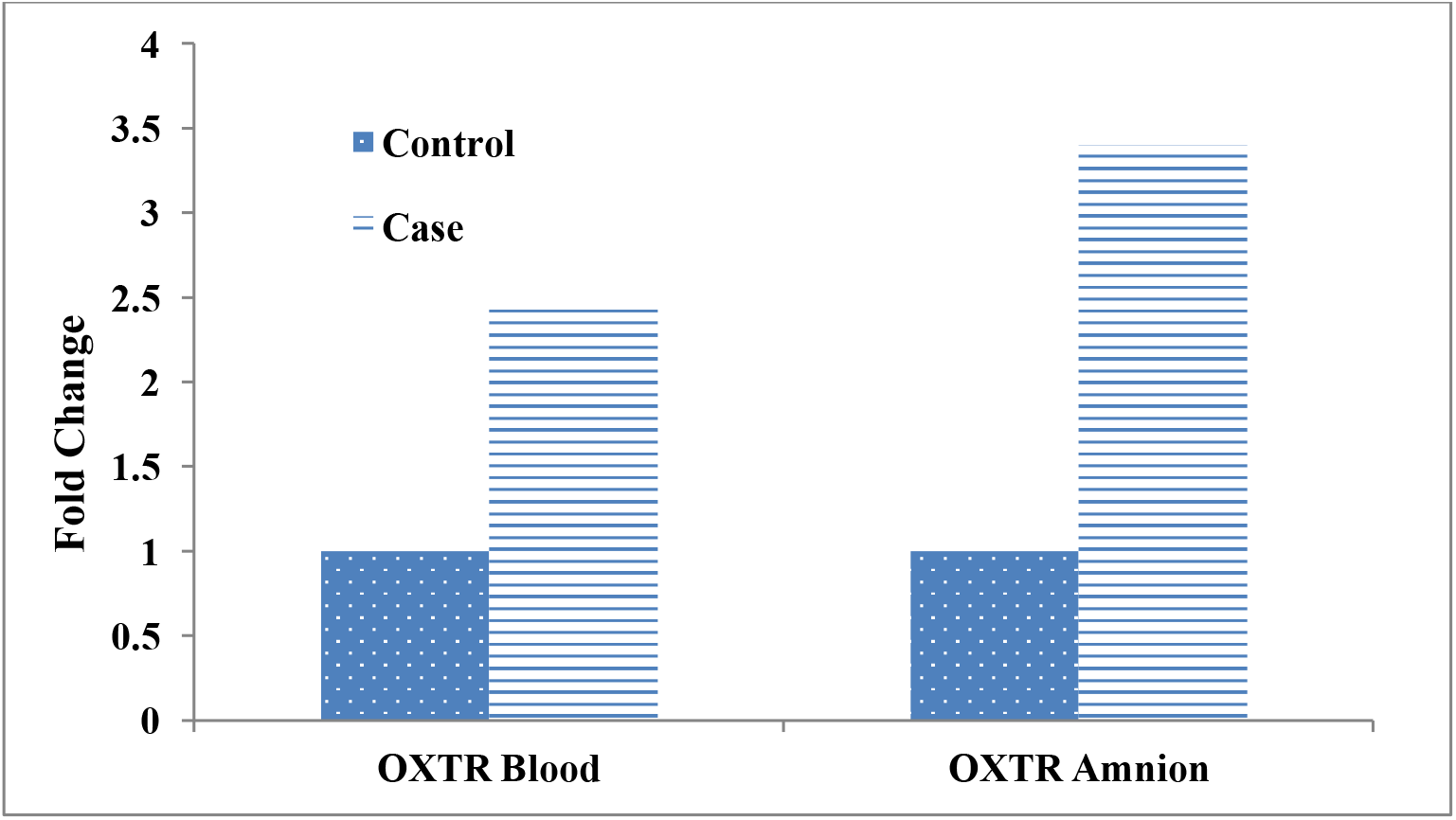
OXTR gene expression in maternal blood and Amnion expressed in terms of Fold Change.

**Table 4:** Correlation between maternal serum oxytocin levels with OXTR gene expression in maternal blood and amnion and with POG, B.W, and placental weight in *preterm birth* cases (n=70)

| Parameter |  | Maternal<br>Serum OXT<br>levels | OXTR<br>maternal<br>blood ΔCq | OXTR<br>amniotic<br>ΔCq | POG | B.WT. (Kg) | Placenta<br>WT.<br>(gms) |
| --- | --- | --- | --- | --- | --- | --- | --- |
| Maternal<br>Serum OXT<br>levels | r | 1 | 0.074 | 0.027 | -0.008 | 0.063 | 0.027 |
|  | p | – | 0.544 | 0.824 | 0.945 | 0.605 | 0.827 |
| OXTR<br>maternal blood<br>ΔCq | r | 0.074 | 1 | -0.116 | 0.048 | -0.130 | -0.145 |
|  | p | 0.544 | – | 0.338 | 0.695 | 0.282 | 0.231 |
| OXTR<br>Amnion ΔCq | r | 0.027 | - 0.116 | 1 | 0.074 | 0.001 | -0.022 |
|  | P | 0.824 | 0.338 | – | 0.543 | 0.995 | 0.857 |

\*p value < 0.05 is significant
r = Pearson correlation coefficient

**Table 5:**
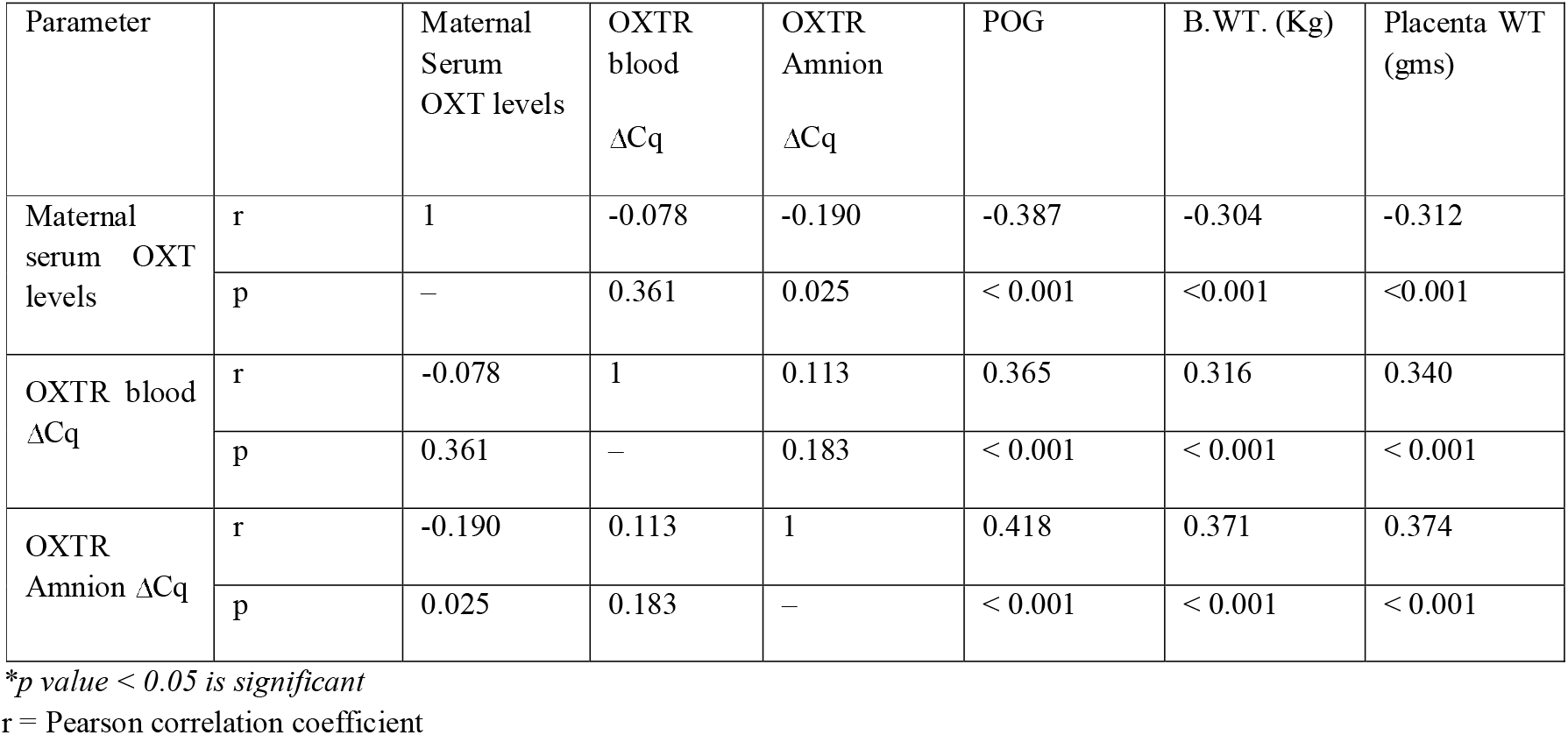
Correlation between maternal serum oxytocin levels with OXTR gene expression in maternal blood and amnion and with POG, B.W, and placental weight in *preterm birth cases & term controls combined (n=140)*.

| Parameter | | Maternal Serum OXT levels | OXTR blood $\Delta Cq$ | OXTR Amnion $\Delta Cq$ | POG | B.WT. (Kg) | Placenta WT (gms) |
| --- | --- | --- | --- | --- | --- | --- | --- |
| Maternal serum OXT levels | r | 1 | -0.078 | -0.190 | -0.387 | -0.304 | -0.312 |
|  | p | – | 0.361 | 0.025 | < 0.001 | <0.001 | <0.001 |
| OXTR blood $\Delta Cq$ | r | -0.078 | 1 | 0.113 | 0.365 | 0.316 | 0.340 |
|  | p | 0.361 | – | 0.183 | < 0.001 | < 0.001 | < 0.001 |
| OXTR Amnion $\Delta Cq$ | r | -0.190 | 0.113 | 1 | 0.418 | 0.371 | 0.374 |
|  | p | 0.025 | 0.183 | – | < 0.001 | < 0.001 | < 0.001 |

\*p value < 0.05 is significant
r = Pearson correlation coefficient

**Fig. 2:**
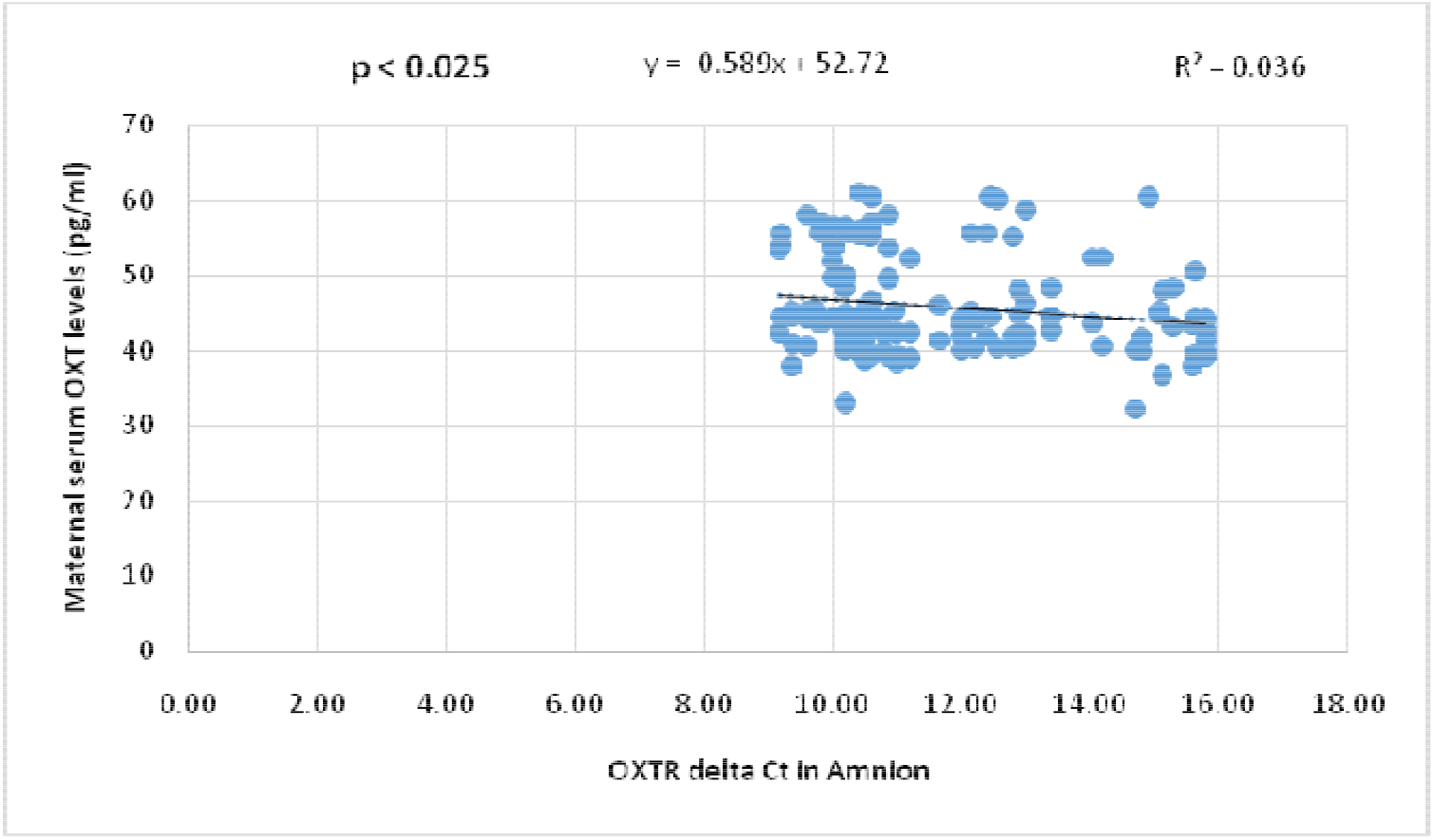
Scatter dot plot graph showing negative correlation between OXTR ΔCq amnion and maternal serum OXT levels in the whole study population.

Pearson correlation was used to correlate maternal serum OXT levels, OXTR gene expression in maternal blood and amnion with the period of gestation, birth weight and placental weight. A positive correlation was seen between OXTR ΔCq blood and OXTR ΔCq amnion with birth weight and placental weight (i.e. negative correlation between OXTR gene expression in blood and amnion with birth weight and placental weight) in PTL cases (Table 4). So, as the OXTR expression in blood and amnion increased there was corresponding decrease in POG (i.e. PTB), birth weight and placental weight.

Similarly, negative correlation was seen between maternal serum OXT levels with the period of gestation, birth weight and placental weight in the whole study population (Table 5, Fig.3 & Fig.4). So, as maternal serum OXT levels increased; there is corresponding significant reduction in POG, birth weight, placental weight.

**Fig. 3:**
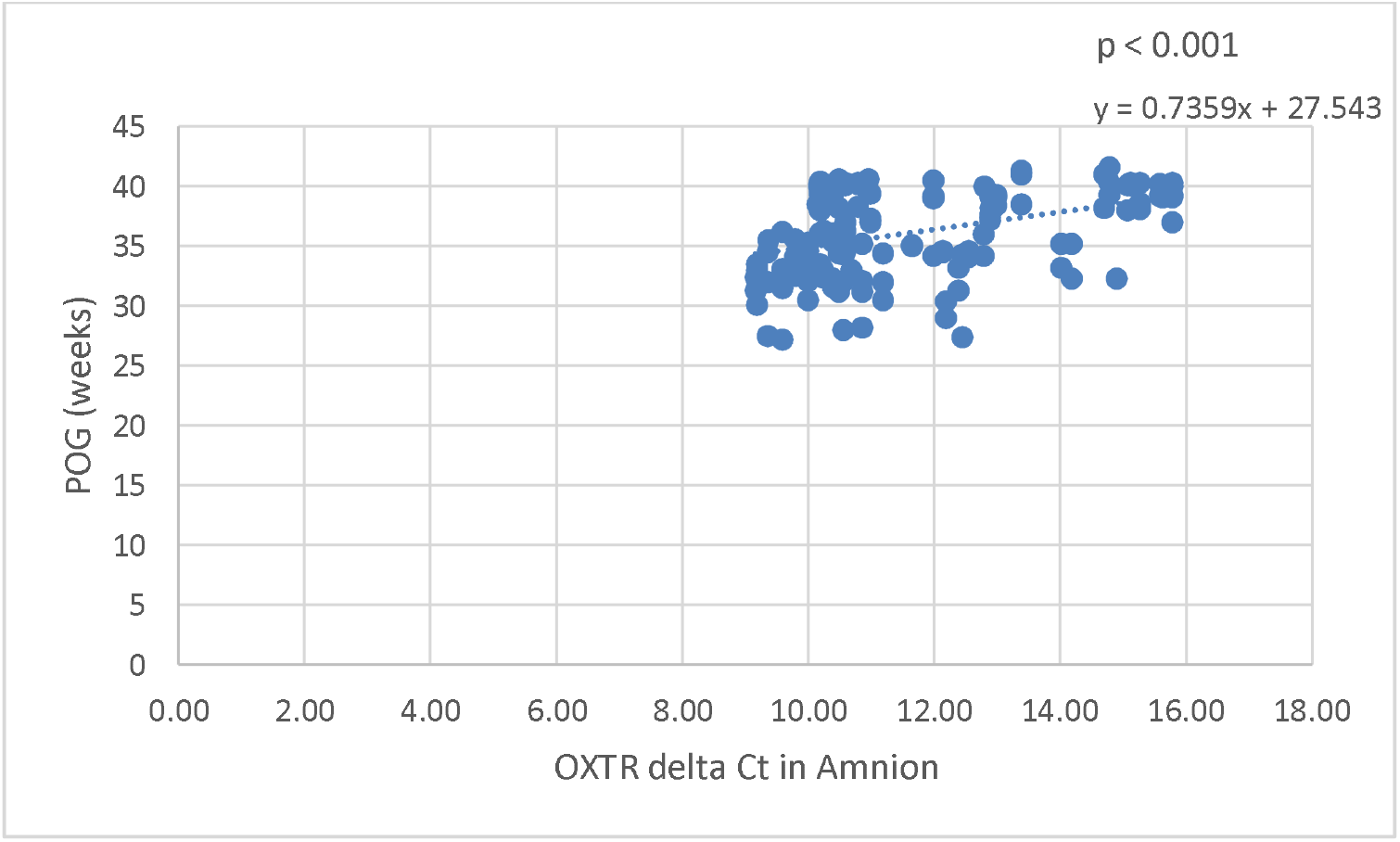
Scatter dot plot graph showing positive correlation between OXTR delta Ct in Amnion and period of gestation in preterm & term subjects combined

**Fig. 4:**
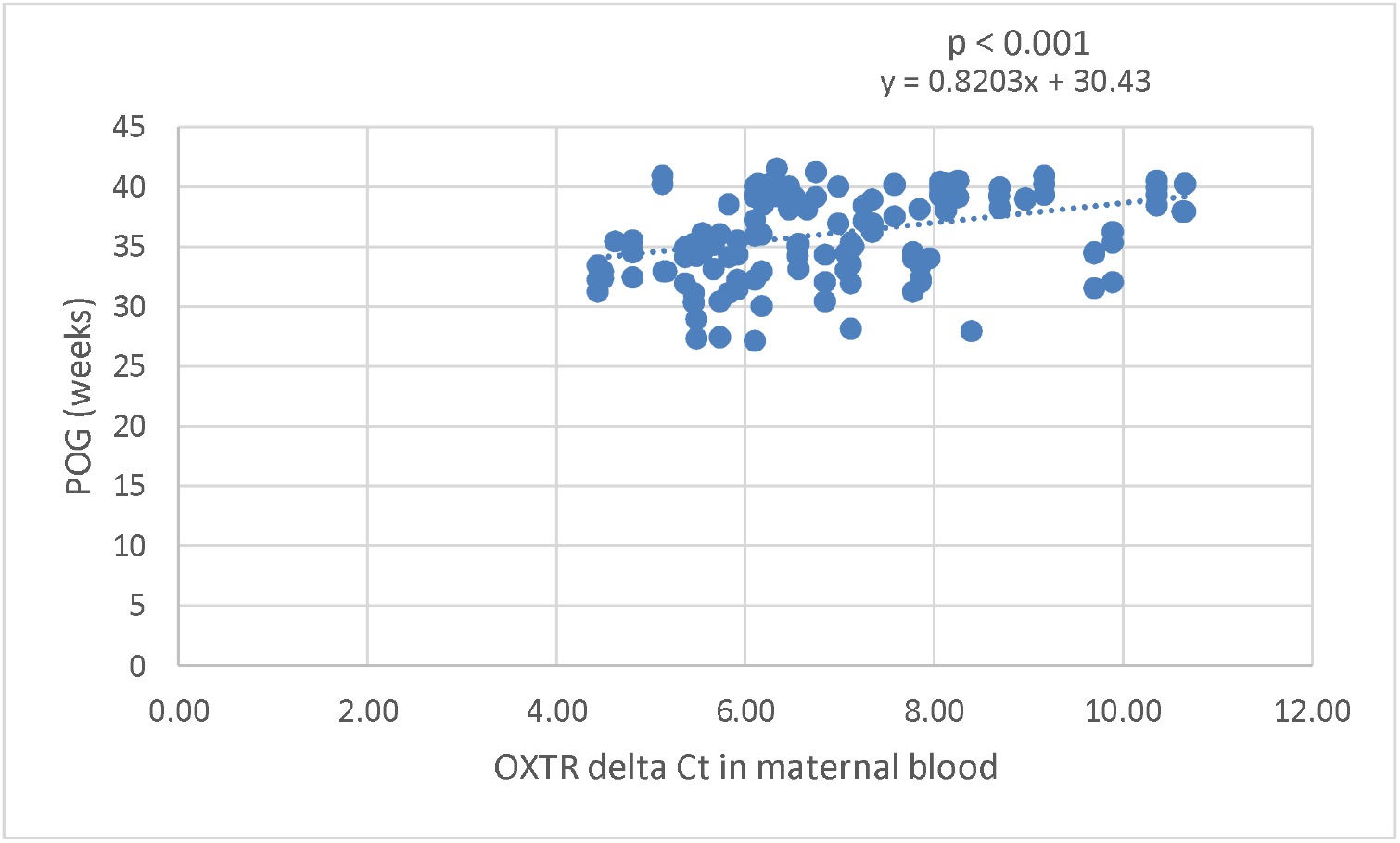
Scatter dot plot graph showing positive correlation between OXTR delta Ct in maternal blood and period of gestation in preterm & term subjects combined

## DISCUSSION

The mean serum OXT level in PTB cases was 48.56 ± 6.97 pg/ml; significantly higher than in controls (43.00 ± 3.96 pg/ml), p<0.001 similar to earlier studies. OXTR gene expression in maternal blood was significantly higher (2.44 folds) in the PTB cases as compared to term controls (p < 0.001). There is also an increase in OXTR mRNA in human amniotic cells up to 3.44 folds in preterm. There was a significant effect of labor process; both in blood and amnion on OXTR mRNA concentrations increasing significantly after the initiation of active phase of labor. The local increase in the estrogen/progesterone ratio in late pregnancy may activate OXT gene transcription and increase the number of OXTRs in myometrium and other gestational tissues. The latter may sensitize the myometrium to any local OXT, thereby leading to increased contractile activity and production of stimulatory PGs. The increased PGs would directly stimulate the myometrium and potentially lead to further increase in production of OXT and its receptor. This coordinated interaction involving OXT, OXTRs and PGs in human fetal membranes could ultimately result in the onset of parturition. OXTRs are present in blood & amnion and show major regulatory changes at the onset of labor whether at term or preterm with increased expression in PTB cases.

*Terzidou et al* [4] showed increased synthesis of PGs in human amnion between 2 and 6 hours following OXT treatment and was associated with increased PGS2 expression. The increased ability of human amnion to produce PGE2in response to OXT treatment suggests a complementary role of the OXT/OXTR system in the activation of human amnion and in the onset of labor. This suggests OXT-PGS autocrine paracrine circuit system in decidua, amnion and myometrium induce and facilitate labor in situ. It makes sense that plasma OXT levels do not alter prior to the onset of labor if this system is mainly regulated within decidua and fetal membranes in situ. *Szukiewicz et al* [8] concluded that upregulation of OXTR within placental trophoblast cells localized close or adherent to uterine wall may play a crucial role in labor with efficient contractile activity leading to vaginal delivery. *Kim et al*^6^ showed that OXT increases the expression of COX-2 and other inflammatory mediators known to be associated with the onset of labor in both the myometrium and amnion via activation of NF-κB and MAPKs. OXT-OXTR interaction leads to Nuclear factor (NF–kβ) activation and subsequent upregulation of PGS, inflammatory chemokines and cytokines that are known to play key role in fetal membrane remodelling, cervical ripening and myometrial activation.

Atosiban is a mixed oxytocin/vasopressin receptor competitive antagonist. It has actions on both the OXTR and vasopressin (V_1a_) receptor. Atosiban has limited bioavailability and requires parenteral administration and hospitalisation. Its low affinity for the OXTR and antagonistic activities at the V_1a_R, has ignited interest in the development of other peptide and nonpeptide antagonists with greater specificity for the OXTR. Further, Atosiban, activates inflammatory pathways in amnion in a way similar to OXT and increases cytokine/chemokine and PG secretion, which may have detrimental effects upon the fetus also in PTL at early gestational ages [9]. It appears from the present study that a molecule with targeted intervention at amnionic OXTR gene may be a promising drug prospect to prevent PTL. The significant reduction in gestational length (PTL) with overexpression of OXTR gene in amnion & blood could be used as a landmark for future studies on formulating preventive strategies for preterm birth.

## CONCLUSION

Significantly higher maternal serum OXT levels seen in PTB cases suggest that the basic regulation for uterine activation was similar between term and PTL. The OXTR expression in blood & amnion are 2.44 folds & 3.44 folds higher respectively in women having PTB. Upregulation of OXTR expression in amnion seems to be crucial in the mechanisms involved in PTL leading to contractile activity and PTB. Amnion thus may be acting as a major site of prostaglandins production on stimulation of OXTRs and linking OXT-PGS autocrine paracrine circuit system to facilitate PTL. Future studies may be planned to unveil these local OXT/OXTR signalling so as to devise better OXTR antagonists preferably targeting amnionic OXTR receptors to prevent PTB.

## Data Availability

All the relevent data pertaining to the study have been provided in the manuscript. Further data will be provided on request.

---

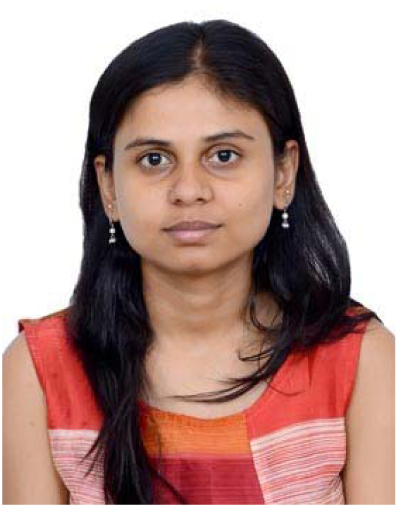

Dr Kumari Anukriti presently working as senior resident in obstetrics & gynaecology department of AIIMS, delhi has completed her MD degree from UCMS &GTB Hospital, delhi in year 2017. She has received Dr. RD PANDIT Research Prize Award for best PG Thesis in 60^th^ AICOG held on 25-29^TH^ January,2017. She has received Silver Medal in 38^th^ annual conference of AOGD for competition paper presentation. Also received GOLD medal for free paper presentation on Bad Obstetric history theme in FOGSI conference in association with AOGD in 2017. She is working in the area of adverse reproductive outcomes such as preterm birth (PTB), fetal growth restriction, and recurrent miscarriages under guidance of Professor Neerja Bhatla in AIIMS Delhi.

